# Measurement equivalence of the SRQ-20 across armed-conflict exposure, sex, and region in Colombia (ENSM 2015)

**DOI:** 10.64898/2026.07.23.26358797

**Authors:** Pedro Vélez-Pardo, Daniela Sánchez Acosta, Nadia Semenova Moratto-Vásquez, Johana Marcela Quintero-Hoyos

## Abstract

**Background:** The SRQ-20 screens common mental distress in low- and middle-income countries, yet its equivalence across groups—especially armed-conflict exposure—is rarely tested with methods that separate true invariance from an underpowered null. We evaluated its psychometric properties and measurement equivalence in Colombian adults.

**Methods:** In 10,865 adults from the 2015 Colombian National Mental Health Survey, we assessed dimensionality, fitted a two-parameter logistic (2PL) model, and tested equivalence across armed-conflict exposure, sex, and region using multiple-group models with purified anchoring and freely estimated group means, separating true distress differences from item bias. Item functioning was equivalence-tested against a *±*0.10 band on signed expected-score differences (SIDS), with test-level differential test functioning (DTF) plus severity-graded and design-weighted sensitivity analyses. Criterion validity used design-weighted ROC against 12-month CIDI diagnoses.

**Results:** The scale was essentially unidimensional (one-factor CFI = .945, rising to .971 with four content-redundant item pairs modelled; explained common variance = .71) and fit the 2PL well, with high conditional reliability at the cut-points (.93–.94). Once true distress differences were separated from item bias, the SRQ-20 was equivalent across armed-conflict exposure (all |SIDS| < .10, maximum .03; net DTF *≈* 0.1 points) and region, holding even among directly victimised adults; sex was partially invariant (three items; DTF *≈* 0.85 points). Design-weighted AUC was .88 (major depression) and .84 (any disorder).

**Conclusions:** The SRQ-20 measures distress equivalently across armed-conflict exposure (including direct victimisation) and region in Colombian adults, supporting exposed–non-exposed comparisons within this population; raw-total sex comparisons carry a small, quantifiable bias. The 20-item form is recommended.

**Highlights:** *(Submit as a separate file; 3–5 points, ≤85 characters each.)*

- The SRQ-20 measures distress equivalently across armed-conflict exposure in Colombia
- Equivalence held after separating true distress differences from item bias
- It held even among directly victimised adults (a severity-graded check)
- Sex was only partially invariant: a ∼0.85-point net test-functioning bias
- Design-weighted criterion validity against CIDI was good (AUC = .88/.84)

## 1. Introduction

Common mental disorders—principally depressive and anxiety syndromes—are leading contributors to the global burden of disease, and their detection in general and primary-care settings depends heavily on brief screening instruments. In low- and middle-income countries (LMICs), the World Health Organization’s Self-Reporting Questionnaire (SRQ-20) has been one of the most extensively deployed screens for non-psychotic psychological distress since its development (Harding et al., 1980; Beusenberg and Orley, 1994), owing to its brevity, free availability, and cross-cultural track record; validation studies span Latin America, sub-Saharan Africa, and South Asia (Mari and Williams, 1986; Scholte et al., 2011; van der Westhuizen et al., 2016; Netsereab et al., 2018), and it remains among the few screening tools with validated performance across diverse LMIC settings (Ali et al., 2016). Despite this ubiquity, the psychometric evidence base for the SRQ-20 remains uneven: many applications rely on classical reliability coefficients and a presumed unidimensional total score (Iacoponi and Mari, 1989; Santos et al., 2009), while comparatively few studies have interrogated its item-level behaviour with item response theory (IRT) or, crucially, tested whether the instrument measures the same construct on the same metric across the sociodemographic and exposure groups that its scores are routinely used to compare (Pendergast et al., 2014; Paraventi et al., 2015; Burnette et al., 2024).

The question of measurement equivalence is not merely technical. Whenever SRQ-20 scores are compared between men and women, across regions, or between populations differentially exposed to adversity, an implicit assumption of measurement invariance is being made—namely, that a given level of underlying distress produces the same expected item responses regardless of group membership (Meredith, 1993; Vandenberg and Lance, 2000; Putnick and Bornstein, 2016). If that assumption fails (differential item functioning, DIF), observed group differences in scores may reflect artefacts of measurement rather than true differences in distress, with direct consequences for epidemiological estimates and resource allocation (Steinberg and Thissen, 2006). In settings marked by prolonged armed conflict, this concern is acute: it is frequently assumed, but rarely tested, that instruments developed and validated in non-conflict contexts behave equivalently among conflict-affected populations (Miller and Rasmussen, 2010; Charlson et al., 2019).

Colombia offers an informative case. The country experienced more than five decades of internal armed conflict, and its 2015 National Mental Health Survey (Encuesta Nacional de Salud Mental, ENSM) was fielded during the peace negotiations, incorporating both the SRQ and a structured diagnostic interview (a reduced Composite International Diagnostic Interview, CIDI; Robins et al., 1988; Kessler and Üstün, 2004) in a nationally representative probability sample (Gómez-Restrepo et al., 2016a), which documented substantial 12-month prevalence of depressive and anxiety disorders among Colombian adults (Gómez-Restrepo et al., 2016b). Conflict-exposed Colombians report substantially more psychological distress, and displacement and cumulative victimisation track with worse mental-health outcomes (Bell et al., 2012; Cuartas Ricaurte et al., 2019; Marroquín Rivera et al., 2020), consistent with the elevated prevalence of mental disorders documented across conflict-affected populations globally (Steel et al., 2009; Charlson et al., 2019; Tamayo-Agudelo and Bell, 2019). Although the SRQ-20 has been applied in specific Colombian subpopulations (Fandiño-Losada et al., 2022), the instrument itself has not been subjected to a national-scale IRT calibration, and its measurement invariance across conflict exposure, sex, and region has not been established. More broadly, whether a major population stressor such as armed conflict alters the *measurement* of distress—how symptoms index the underlying construct—as opposed to simply elevating its *level*—remains an open and consequential question.

The present study addresses these gaps. Using the ENSM 2015 adult sample, we (i) characterise the dimensionality, item-response parameters, model–data fit, and reliability of the SRQ-20; (ii) test measurement invariance and item-level DIF across armed-conflict exposure, sex, and region, using effect-size and equivalence-testing criteria appropriate to a very large sample (Meade, 2010; Nye and Drasgow, 2011; Lakens, 2017), with the DIF re-estimated under purified anchoring and a freely estimated group latent mean so that true group differences in distress are separated from item bias; (iii) establish criterion validity against 12-month CIDI diagnoses, including practical cut-points; and (iv) evaluate the 25-item extension that adds four psychotic-experience items. A moderated psychometric-network analysis is reported in the Supplementary Materials as exploratory, convergent evidence only. Critically, because our primary substantive interest is a claim of *equivalence* across conflict exposure, we treat a non-significant group difference not as proof of invariance but as a hypothesis requiring positive equivalence evidence and adequate statistical power (Lakens et al., 2018).

## 2. Methods

### 2.1 Participants and design

Data came from the 2015 Colombian National Mental Health Survey (ENSM), a nationally representative, multistage probability survey of the non-institutionalised population conducted by the Ministry of Health, Pontificia Universidad Javeriana, and Colciencias (Ministerio de Salud y Protección Social and Colciencias, 2015; Gómez-Restrepo et al., 2016a). The present analyses concern adults aged 18 years and older. The SRQ was administered to 10,870 adults; complete SRQ-20 responses were available for 10,865 (99.95%), who constitute the analytic sample. Sampling expansion weights (FEX) were available; primary sampling units and design strata are not released in the public microdata, a limitation addressed below.

### 2.2 Measures

#### SRQ-20/25

The SRQ comprises 20 dichotomously scored (yes/no) items indexing non-psychotic distress over the preceding 30 days (e.g., headaches, poor sleep, tearfulness, feeling worthless, thought of ending one’s life), plus four psychotic-experience items (SRQ-21–24: persecutory ideation, grandiosity, thought interference, hallucinations) and one neurological item (SRQ-25, convulsions) (Beusenberg and Orley, 1994). Items were coded 1 = symptom present, 0 = absent; the psychiatric analyses use items 1–20 (primary) and 1–24 (extension), excluding the neurological item. The full English item wording is given in Table 1; the SRQ is freely available from the World Health Organization (Beusenberg and Orley, 1994).

**Table 1.** Item wording (WHO), 30-day endorsement, corrected item–total correlation, and 2PL discrimination (a) and difficulty (b), SRQ-20.

| Item | SRQ item (WHO) | Endorsement | item–total |  |  |
| --- | --- | --- | --- | --- | --- |
|  |  | (%) | r | a | b |
| 1 | Do you often have headaches? | 21.6 | .35 | 1.10 | 1.44 |
| 2 | Is your appetite poor? | 9.4 | .37 | 1.43 | 2.08 |
| 3 | Do you sleep badly? | 16.0 | .46 | 1.66 | 1.44 |
| 4 | Are you easily frightened? | 23.7 | .42 | 1.39 | 1.14 |
| 5 | Do your hands shake? | 6.3 | .33 | 1.48 | 2.38 |
| 6 | Do you feel nervous, tense or worried? | 15.8 | .58 | 2.58 | 1.22 |
| 7 | Is your digestion poor? | 10.1 | .36 | 1.35 | 2.08 |
| 8 | Do you have trouble thinking clearly? | 9.2 | .48 | 2.12 | 1.72 |
| 9 | Do you feel unhappy? | 16.4 | .58 | 2.61 | 1.18 |
| 10 | Do you cry more than usual? | 11.1 | .48 | 2.03 | 1.62 |
| 11 | Do you find it difficult to enjoy your daily activities? | 6.8 | .52 | 2.79 | 1.76 |
| 12 | Do you find it difficult to make decisions? | 9.7 | .49 | 2.11 | 1.69 |
| 13 | Is your daily work suffering? | 5.3 | .43 | 2.27 | 2.04 |
| 14 | Are you unable to play a useful part in life? | 2.5 | .28 | 1.81 | 2.76 |
| 15 | Have you lost interest in things? | 8.7 | .51 | 2.37 | 1.69 |
| 16 | Do you feel that you are a worthless person? | 3.2 | .31 | 1.70 | 2.70 |
| 17 | Has the thought of ending your life been on your mind? | 6.5 | .35 | 1.54 | 2.30 |
| 18 | Do you feel tired all the time? | 10.1 | .52 | 2.38 | 1.59 |
| 19 | Do you have uncomfortable feelings in your stomach? | 8.1 | .44 | 1.80 | 1.95 |
| 20 | Are you easily tired? | 17.6 | .53 | 2.14 | 1.21 |

#### CIDI diagnoses

Twelve-month diagnoses of major depression and of any assessed mental disorder were derived from the ENSM’s structured diagnostic module—a reduced form of the WHO Composite International Diagnostic Interview (Robins et al., 1988; Kessler and Üstün, 2004)—and used as criterion standards (analytic n = 10,399 and 10,404; prevalence 2.0% and 3.8%).

#### Grouping variables

Armed-conflict exposure was operationalised from the traumatic-events checklist (module M13b) as endorsement of victimisation by, witnessing of, or vicarious exposure to armed-conflict events (enforced disappearance, torture, kidnapping, forced recruitment, massacres, landmines, armed confrontations, or conflict-related sexual violence). Sex (men/women) and region (five macro-regions) were taken from the household and person modules.

### 2.3 Statistical analysis

#### Dimensionality

Because the items are dichotomous, dimensionality and CFA used tetrachoric correlations and the mean- and variance-adjusted weighted least squares (WLSMV) estimator (Flora and Curran, 2004; Rhemtulla et al., 2012; Li, 2016). We evaluated Horn’s parallel analysis (Horn, 1965; Timmerman and Lorenzo-Seva, 2011) and the ratio of the first to second eigenvalue; bifactor-based dimensionality indices—explained common variance (ECV), *ω*-hierarchical, and the proportion of uncontaminated correlations (PUC)—computed from a Schmid–Leiman transformation of an exploratory bifactor solution (Reise, 2012; Rodriguez et al., 2016a, 2016b); and confirmatory one-factor, correlated two-factor (somatic vs psychological), and bifactor models (fit thresholds: CFI/TLI *≥* .95 good, RMSEA *≤* .06 good, SRMR *≤* .08 good; Hu and Bentler, 1999). Because a few content-redundant item pairs can inflate apparent multidimensionality, we additionally fitted a one-factor model with correlated residuals for the locally dependent pairs (a testlet parameterisation; see Section 3.4) and compared its fit with the plain one-factor model.

#### Item-response model

A two-parameter logistic (2PL) model estimated item discrimination (a), difficulty (b), and the test-information function by marginal maximum likelihood in mirt (Chalmers, 2012). The 2PL was compared against a one-parameter (equal-slope) model by likelihood-ratio test and information criteria (AIC, BIC) to justify allowing discriminations to vary (Embretson and Reise, 2000). Global model–data fit was assessed with the limited-information M2* statistic and its associated RMSEA and SRMSR (Maydeu-Olivares and Joe, 2005; Maydeu-Olivares, 2013); item-level fit with the S-X^2^ statistic and its RMSEA (Orlando and Thissen, 2000); and the local-independence assumption with Yen’s Q3 residual correlations, flagging pairs exceeding the mean Q3 by more than 0.20 (Yen, 1984; Christensen et al., 2017). Reliability was indexed by Cronbach’s *α* (Cronbach, 1951), McDonald’s *ω* and *ω*-hierarchical (McDonald, 1999; Zinbarg et al., 2005; Revelle and Zinbarg, 2009), IRT marginal and empirical reliability, and **conditional reliability and standard error at the screening cut-points**, obtained by mapping cut-scores to *θ* via the test characteristic curve.

#### Measurement equivalence

For each grouping variable we fitted a hierarchy of multiple-group 2PL models—configural (all parameters free), metric (equal discriminations), and scalar (equal discriminations and difficulties)—comparing adjacent models with the change in the C2-based comparative fit index (ΔCFI *≤* .010 indicating invariance; Cheung and Rensvold, 2002; Chen, 2007; Putnick and Bornstein, 2016). Because ΔCFI is insensitive under the group-size imbalance here (up to ∼12:1) and because “failure to reject” is not evidence of equivalence, we additionally performed **equivalence testing on item-level DIF effect sizes** (Lakens, 2017; Lakens et al., 2018).

The essential step is to separate genuine item bias from a group difference in the latent trait (*impact*): if a more-distressed focal group’s latent mean is fixed at zero, that difference is absorbed into the item parameters as spurious, uniformly-signed DIF. We therefore used a **purified-anchor procedure with a freely estimated group latent mean and variance**. An initial configural model ranked items by their absolute signed expected-score difference (SIDS; Steinberg and Thissen, 2006; Meade, 2010; via mirt::empirical_ES); the four most invariant items served as anchors; and the model was refitted with the focal group’s mean and variance freely estimated (Reise et al., 1993; Millsap, 2011; Tay et al., 2015). SIDS were recomputed from this purified model, and the estimated focal-group latent mean is reported as the impact. An item met equivalence when its SIDS fell within a pre-specified band of |SIDS| < 0.10—less than half a point on the 0–20 scale, corresponding to under a ten-percentage-point difference in the probability of endorsing the item at equal latent distress (Nye and Drasgow, 2011). Each SIDS was bootstrapped (stratified resampling) for a 95% CI; because the public microdata omit the sampling design (Section 4.1), we also report intervals inflated for a plausible design effect (DEFF *≈* 2). As a detectability check—not a formal equivalence-power calculation—the smallest item DIF the design could resolve was approximated as 2.8 *×* bootstrap SE.

Where DIF exceeded the band we retained a **partial-invariance** model (flagged items free, remainder anchored) and quantified the net scale-level consequence via **differential test functioning (DTF)** at the observed group trait distributions: the signed test difference (STDS, the sum of the signed item expected-score differences) and the unsigned test difference (UTDS, the sum of the *absolute* item expected-score differences), following Meade (2010); the DTF was bootstrapped for a 95% CI and interpreted against the raw-score metric. Two robustness analyses addressed the equivalence claim directly. A **severity-graded (dose–response) analysis** repeated the conflict DIF using only adults reporting *direct* victimisation versus the non-exposed, testing whether equivalence survives a cleaner, more severe exposure contrast (which also bounds the effect of non-differential exposure misclassification). A **design-weighted sensitivity analysis** re-estimated all item parameters with the expansion weight (FEX; mirt survey.weights) to confirm that discriminations, difficulties, and DIF are stable under weighting. Region (five groups) was assessed by the ΔCFI hierarchy and, because the SIDS index is defined for two-group contrasts, by computing purified SIDS for each region against the pooled remainder and taking the maximum across the five contrasts.

#### Criterion validity

Design-weighted (FEX) ROC analysis of the SRQ-20 sum against each CIDI outcome, reporting the area under the curve (AUC; Hanley and McNeil, 1982; DeLong et al., 1988) with region-stratified bootstrap CIs (WeightedROC); a **content-overlap sensitivity analysis** re-estimating the AUC from the 10 SRQ items that do not overlap conceptually with depression criteria; weighted operating characteristics (sensitivity, specificity, predictive values) at conventional cut-points, interpreted in light of the strong dependence of predictive values on prevalence (Trevethan, 2017; Leeflang et al., 2013); and **.632-bootstrap** validation of the Youden-optimal cut-point (Youden, 1950), with the cut re-selected within each bootstrap replicate, to correct in-sample optimism (Efron and Tibshirani, 1997; Harrell et al., 1996; Steyerberg et al., 2001).

#### Convergent (exploratory) evidence

A moderated network model (mixed graphical model; mgm) with conflict exposure moderating all symptom edges, and an accompanying network-comparison test, are reported in the Supplementary Materials as *exploratory* evidence only; they are underpowered by the small exposed subsample and do not carry the equivalence claim.

#### Software

Analyses used R (R Core Team, 2024): psych (tetrachoric dimensionality, reliability; Revelle, 2026), lavaan (CFA; Rosseel, 2012) with semTools (invariance helpers; Jorgensen et al., 2026), mirt (IRT, model and item fit, local dependence, multiple-group DIF, empirical effect sizes; Chalmers, 2012), WeightedROC (design-weighted ROC; Hocking, 2020), and pROC (Robin et al., 2011). The exploratory network analyses used mgm, bootnet, NetworkComparisonTest, IsingFit, and qgraph (cited in the Supplementary Materials).

## 3. Results

### 3.1 Item descriptives and factorability

Endorsement ranged from 2.5% (unable to play a useful role) to 23.7% (easily frightened), with headache (21.6%), tiredness (17.6%), sadness (16.4%), and poor sleep (16.0%) also common, and the severity items (worthlessness 3.2%, difficulty working 5.3%) rare—an expected pattern for a distress screen in the general population (Table 1). The Kaiser–Meyer–Olkin measure was .914 (meritorious) and Bartlett’s test of sphericity was significant (p < .001), confirming factorability.

### 3.2 Dimensionality

The SRQ-20 is best treated as essentially unidimensional, though the evidence is a matter of degree rather than a clean single-factor fit. Tetrachoric eigenvalues were 10.14, 1.38, and 1.05 (first-to-second ratio 7.4). A Schmid–Leiman transformation of an exploratory bifactor solution yielded a general factor accounting for ECV = .71 of the common variance, with *ω*-hierarchical = .79 and a proportion of uncontaminated correlations PUC = .70; at this moderate PUC, an ECV of .71 with *ω*-hierarchical near .80 indicates that a general factor dominates and that total scores reflect largely a single source of variance (Reise, 2012; Rodriguez et al., 2016a). The confirmatory one-factor model was borderline on one index (CFI = .945, TLI = .939, RMSEA = .043, SRMR = .066): the correlated two-factor (somatic/psychological) model fit marginally better (CFI = .953, RMSEA = .040), but its factors correlated **r = .88**, indicating near-redundancy rather than two separable dimensions. A confirmatory bifactor model also fit well (CFI = .972, RMSEA = .033), but with a strongly dominant general factor and weak specific factors—precisely the configuration the bifactor indices above quantify. Parallel analysis nominally indicated eight factors, but this reflects the well-documented over-extraction of parallel analysis with dichotomous items in very large samples (Timmerman and Lorenzo-Seva, 2011).

The apparent multidimensionality is largely an artefact of a few content-redundant item pairs (Section 3.4). Adding correlated residuals for the four locally dependent pairs to the one-factor model (a testlet parameterisation) raised fit to good on every index (CFI .945 *→* .971, TLI .939 *→* .967, RMSEA .043 *→* .032, SRMR .066 *→* .053), closing most of the gap to the two-factor model without invoking a second substantive dimension. The eigenvalue ratio, the bifactor indices, and the testlet result therefore converge on a single general distress dimension with minor, content-driven local dependence—consistent with the resolution of apparent multidimensionality in comparable health-outcome measures (Reise et al., 2007). We treat the total score as reflecting one general distress factor, while noting that any narrow somatic or affective subscale would need to model these dependencies (Table 1; Table 2; Figure 1; Supplementary Tables S1–S2).

**Figure 1.**
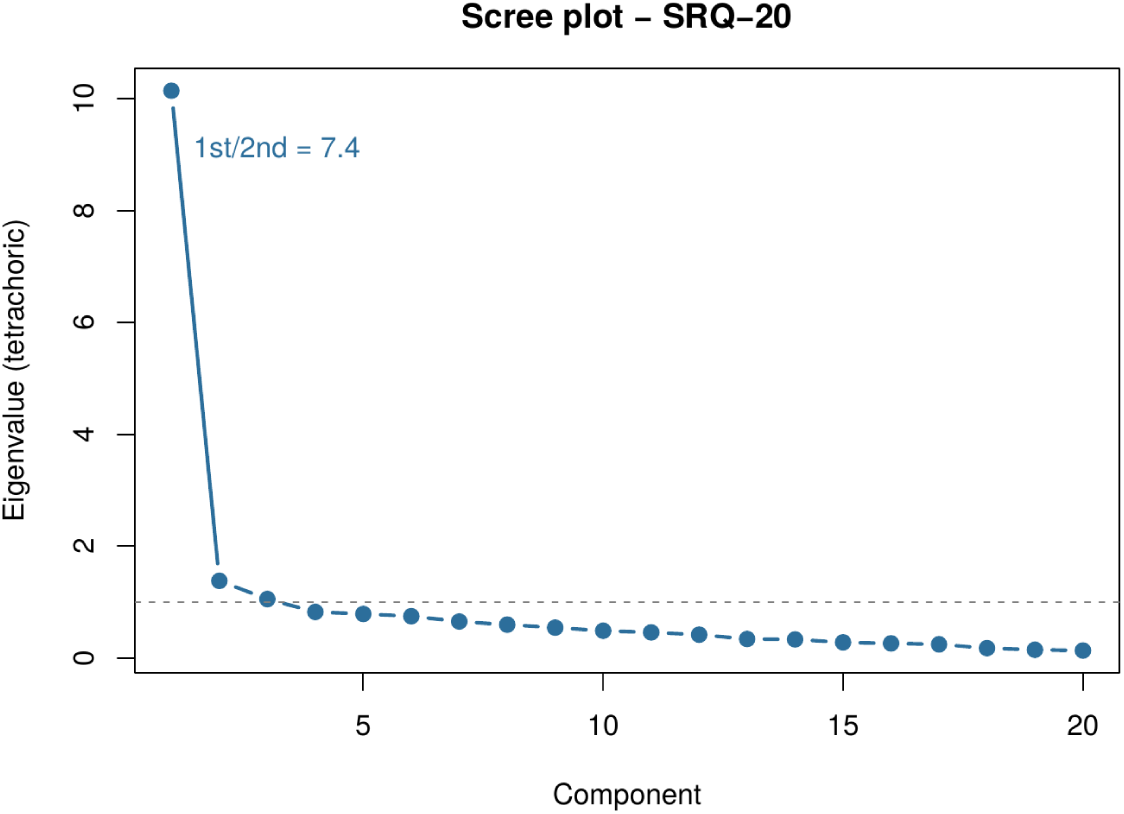
Scree plot of tetrachoric eigenvalues, SRQ-20.

**Table 2.** Dimensionality model comparison (CFA, WLSMV). ECV, *ω*-hierarchical and PUC come from the Schmid–Leiman solution and are reported on a separate row. r_FF = inter-factor correlation.

| Model | CFI | TLI | RMSEA | SRMR | r_FF | ECV | $\omega_h$ | PUC |
| --- | --- | --- | --- | --- | --- | --- | --- | --- |
| One-factor | .945 | .939 | .043 | .066 | — | — | — | — |
| Correlated two-factor | .953 | .947 | .040 | .060 | .88 | — | — | — |
| One-factor + 4 residuals (testlet) | .971 | .967 | .032 | .053 | — | — | — | — |
| Bifactor (CFA) | .972 | .964 | .033 | .049 | — | — | — | — |
| Bifactor indices (Schmid–Leiman) | — | — | — | — | — | .71 | .79 | .70 |

### 3.3 Item-response model: parameters, fit, and information

Allowing discriminations to vary was warranted: the 2PL fit far better than a one-parameter (equal-slope) model (likelihood-ratio *χ*^2^(19) = 1,165, p < .001; AIC 116,579 vs 117,706; BIC 116,871 vs 117,859, ΔBIC *≈* 988 favouring the 2PL). The 2PL showed good global model–data fit: although the limited-information M2* was significant given the sample size (M2*(170) = 3,577, p < .001), the approximate-fit indices were good (RMSEA = .043, 90% CI [.042, .044]; SRMSR = .049; CFI = .971; TLI = .968) (Maydeu-Olivares and Joe, 2005; Maydeu-Olivares, 2013). Item-level fit (S-X^2^) was adequate in magnitude for all 20 items: fifteen items were non-significant, and although five (poor digestion, cry more, worthless, tired all the time, easily tired) reached p < .05, their fit RMSEA values were *≤* .012—negligible misfit reflecting the hypersensitivity of significance tests at n *≈* 10,900 rather than substantive model failure (Orlando and Thissen, 2000) (Supplementary Table S5).

Item discriminations were all acceptable to high (a = 1.10–2.79); the most discriminating items were affective—anhedonia (a = 2.79), sadness (2.61), nervousness (2.58), and tiredness (2.38)—whereas isolated somatic items discriminated least (headache 1.10, indigestion 1.35). All item difficulties were positive (b = 1.1–2.8), so the scale carries most of its information above the population mean: the test-information function peaked at *θ* = 1.64 (information = 17.0, conditional reliability = .94) and declined sharply below the mean. Consequently, IRT marginal reliability was modest (.65) and empirical reliability was .69, well below the classical coefficients (*α* = .852, *ω* = .856; average inter-item r = .227). This is not a defect but a design feature of a screen: the conventional cut-points map to *θ ≈* 1.1 (*≥*5), 1.4 (*≥*7), and 1.5 (*≥*8)—precisely the region of peak information—where **conditional reliability was .93–.94** (SE 0.24–0.27). The SRQ-20 is thus imprecise only where precision is least needed (very low distress) and highly precise at the decision thresholds (item parameters in Table 1; Figure 2).

**Figure 2.**
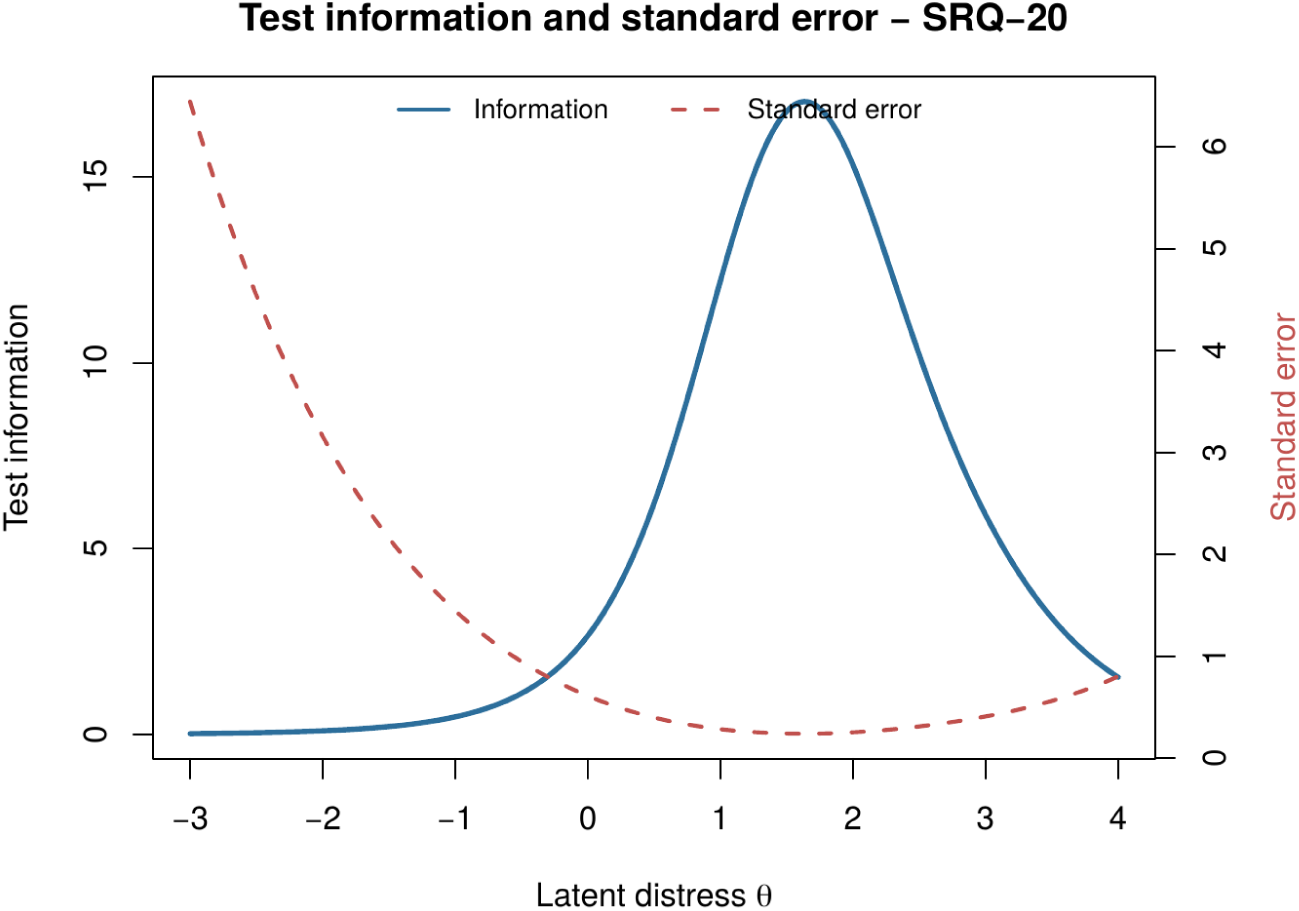
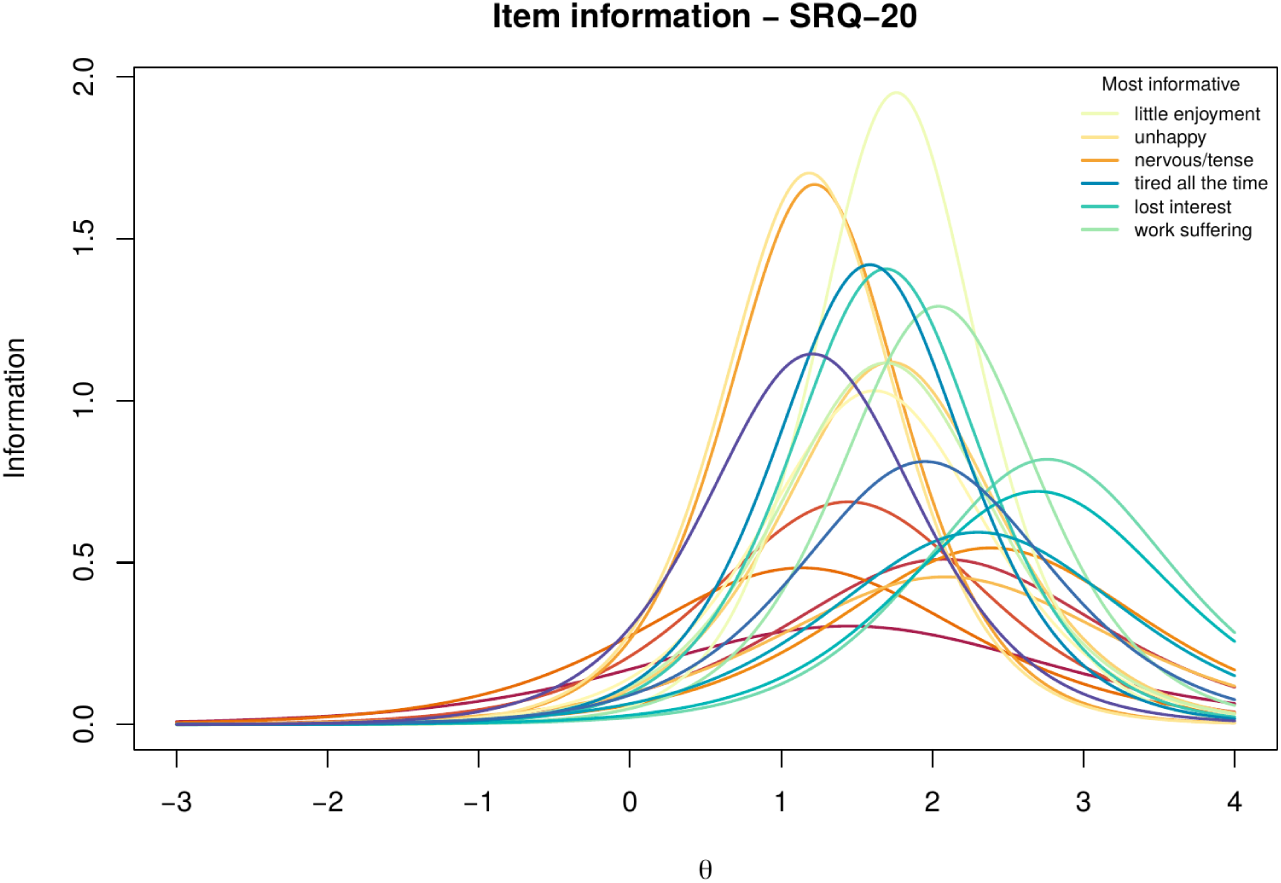
(a) Test information and standard error across the latent distress continuum; (b) item information curves, SRQ-20.

### 3.4 Local dependence

The local-independence assumption held well. Across the 190 item pairs the mean Yen’s Q3 residual correlation was -.042 (the expected slight negative bias) and the maximum absolute value was .28. Applying the Christensen et al. (2017) criterion (|Q3 - mean Q3| > 0.20), only four pairs showed local dependence, and all four were content-redundant: poor digestion–stomach discomfort (Q3 = .26), tired all the time–easily tired (.22), difficulty working–little enjoyment (.28), and worthless–no useful role (.17) (Yen, 1984). This mild, content-driven dependence is consistent with essential unidimensionality (the general factor dominates; ECV = .71) and explains both the parallel-analysis over-extraction and the marginal fit gain of the correlated two-factor model; it does not warrant modelling a second dimension and had no material effect on the item parameters (Supplementary Table S6).

### 3.5 Measurement equivalence across armed-conflict exposure

The SRQ-20 was equivalent across exposed (n = 838) and non-exposed (n = 10,027) adults once true group differences in distress were separated from item bias. The scalar model did not degrade fit (ΔCFI = .0001; LRT p = .22). Tellingly, the configural analysis initially returned all 20 items with same-signed SIDS (net -0.6 points)—the classic signature of a group latent-mean difference leaking into the item parameters rather than genuine DIF. Freeing the focal-group latent mean confirmed exactly this: exposed adults were more distressed (impact = +0.29 SD), and once that impact was modelled the uniform pattern dissolved into small, mixed-signed residuals (13 positive, 3 negative). In the purified model no item approached the equivalence band—the maximum |SIDS| was **.033** (suicidal ideation)—and the smallest DIF the design could resolve was *≈* .03, so the null reflects detectable invariance rather than low power (Lakens et al., 2018). The net differential test functioning was correspondingly small (signed DTF = +0.12, unsigned DTF = 0.18 on the 0–20 scale); unlike the configural result, the signed and unsigned values now diverge, as expected when residual differences are genuine noise rather than systematic bias (Meade, 2010).

Two checks reinforce this conclusion. First, equivalence survived a severity gradient: restricting the exposed group to adults reporting *direct* victimisation (n = 286 vs 10,027 non-exposed)—a cleaner, more severe contrast that also bounds misclassification attenuation—raised the impact to +0.43 SD yet left DIF within the band (maximum |SIDS| = .071, no item *≥* .10; signed/unsigned DTF = 0.27/0.35). Equivalence thus holds, and if anything strengthens, as exposure severity increases. Second, item parameters were essentially unchanged when estimated with the design weights (discrimination and difficulty correlations r = .99; maximum |Δa| = .15), so the DIF results are not an artefact of ignoring the sampling weights (Table 3; Supplementary Table S3; Figure 3a).

**Figure 3.**
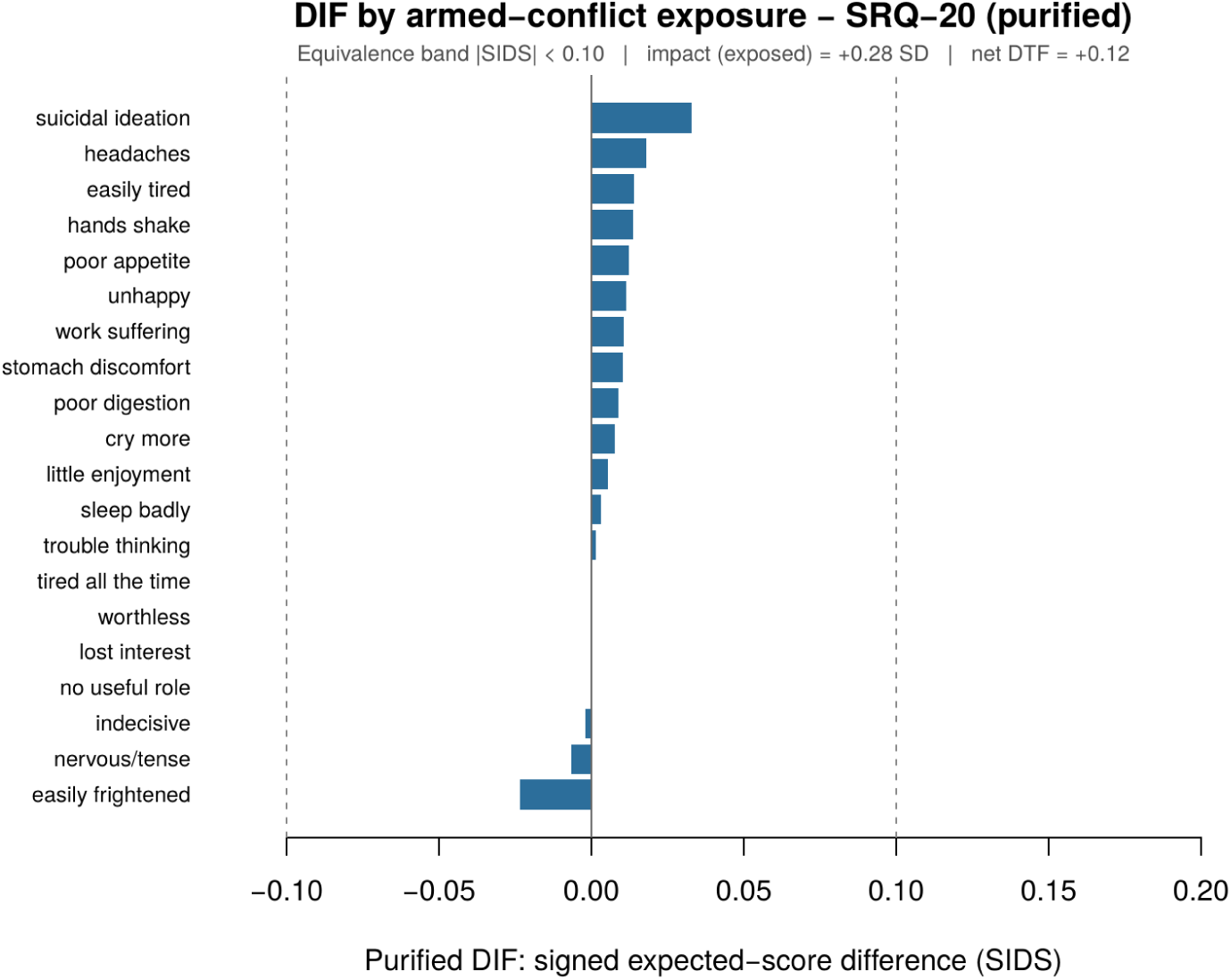
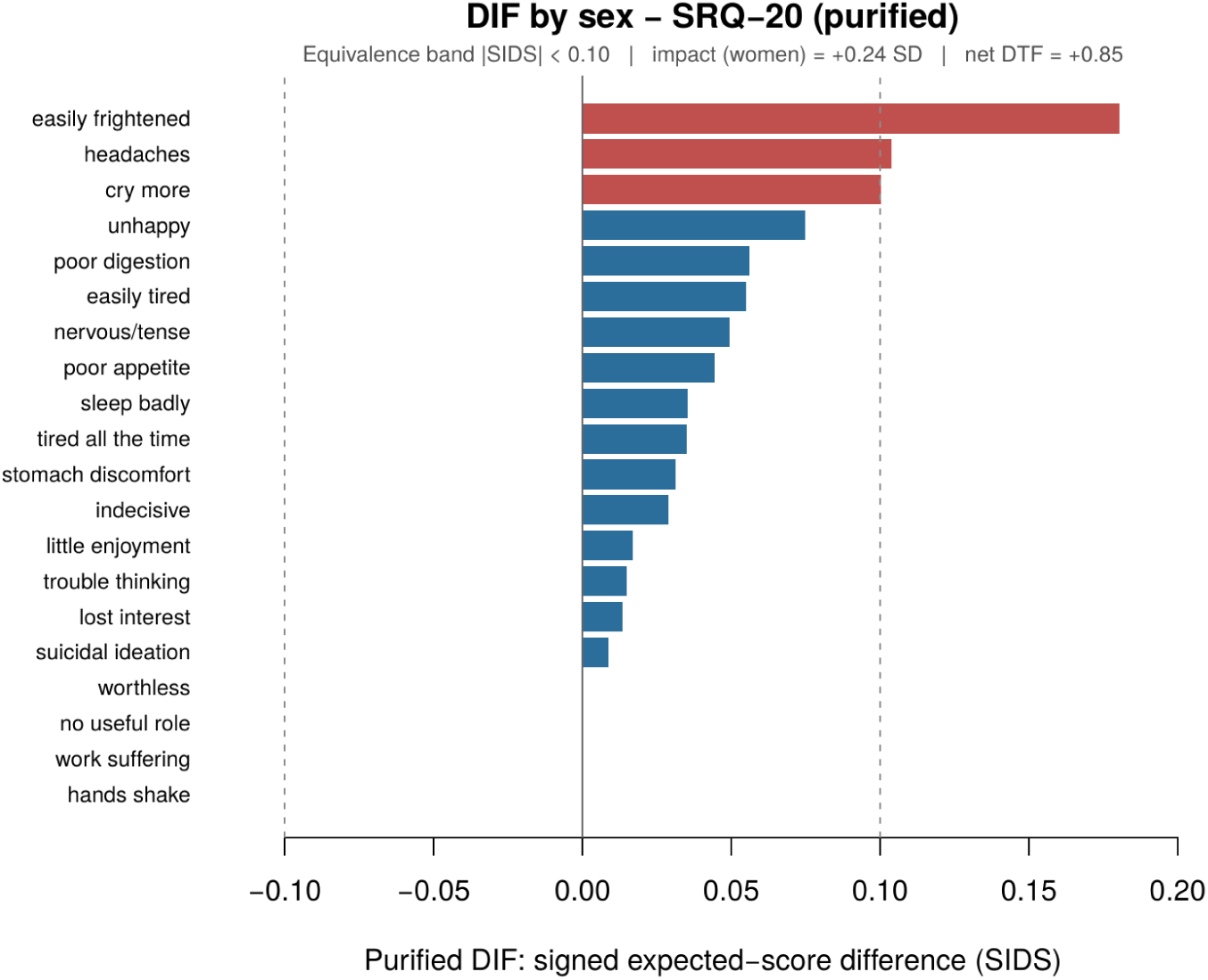
Purified item-level DIF (signed expected-score differences, SIDS) by (a) armed-conflict exposure and (b) sex. Dashed lines mark the equivalence band |SIDS| < 0.10.

**Table 3.** Measurement equivalence by conflict exposure, sex, and region: multigroup fit (ΔCFI) and purified item-level DIF (impact = focal-group latent mean; max |SIDS|; items outside the *±*0.10 band; net signed/unsigned DTF).

| Group | k | CFI<br>conf. | CFI<br>scalar | $\Delta CFI$ | Impact<br>(SD) | max<br>$\vert SIDS\vert $ | N $\geq .10$ | DTF<br>s/u | Conclusion |
| --- | --- | --- | --- | --- | --- | --- | --- | --- | --- |
| Conflict<br>expo-<br>sure | 2 | .971 | .971 | .0001 | +0.28 | .033 | 0 | 0.12 /<br>0.18 | Equivalent |
| Direct<br>victimi-<br>sation<br>(dose-response) | 2 | — | — | — | +0.43 | .071 | 0 | 0.27 /<br>0.35 | Equivalent<br>(sever-<br>ity<br>check) |
| Sex | 2 | .972 | .963 | .0083 | +0.24 | .181 | 3 | 0.85 /<br>0.85 | Partial<br>(3<br>items) |
| Region | 5 | .970 | .968 | .0019 | — | .046 | 0 | — | Equivalent |

### 3.6 Invariance across region; partial invariance across sex

Region (five macro-regions; n = 1,726–2,577) was invariant. The ΔCFI across the five groups was negligible (.002), and purified item-level DIF confirmed it: the maximum |SIDS| across all five region-versus-rest contrasts was .046, well inside the equivalence band.

**Sex was partially invariant**, and it illustrates cleanly why the two invariance criteria can diverge. By the ΔCFI rule, scalar invariance was *retained* (ΔCFI = .0083 *≤* .010); by the more sensitive item-level equivalence test, it was *rejected*. After separating impact from bias—women were the more distressed group (impact = +0.25 SD)—three items still exceeded the band, in every case women endorsing the symptom more at equal latent distress: easily frightened (SIDS = .18), headaches (.10), and crying more (.10); the next-largest item, feeling unhappy (.08), fell below the band. A partial-invariance model freeing these three items fit well. The net differential test functioning was +0.85 points on the 0–20 scale (signed and unsigned DTF both 0.85, since the item differences share direction); that is, at equal latent distress a woman’s expected SRQ-20 total exceeds a man’s by about eight-tenths of a point. This is genuine bias, not impact—it persists after the latent-mean difference is modelled—and it is consistent with the gendered symptom reporting documented for the SRQ elsewhere (Paraventi et al., 2015; Burnette et al., 2024). Because the bias is systematic and same-signed, near the screening thresholds (*θ ≈* 1.3) it can lift individual women across a cut-point: raw-total sex comparisons should therefore be interpreted with an adjustment of about one point (the estimated 0.85-point differential) to women’s scores, or made on the latent metric, rather than taken at face value (Supplementary Table S4; Figure 3b).

### 3.7 Criterion validity

Design-weighted AUC was .877 (95% CI [.845, .900]) for major depression and .836 ([.807, .859]) for any disorder—good discrimination, though bounded by the criterion (below). Content overlap with the criterion inflated discrimination only modestly: the 10 items that do not overlap conceptually with depression criteria still achieved AUC .835 and .799 (a *≈*.04 reduction), so the SRQ’s validity is not merely an artefact of shared content—though, because screen and criterion come from the same self-report interview, some shared-method variance remains. The Youden-optimal cut-point was stable under .632-bootstrap correction with the cut re-selected in each replicate (major depression: sensitivity .76 *→* .75, specificity .79 *→* .79), indicating little in-sample optimism.

Weighted operating characteristics traded sensitivity for specificity across the conventional range: for major depression, *≥*5 gave sensitivity/specificity .71/.86, *≥*7 gave .61/.92, and *≥*8 gave .52/.94; for any disorder, the corresponding values were .60/.87, .49/.93, and .42/.95. Two cautions temper these figures. First, sensitivity is modest even at the lowest cut: *≥*5 misses roughly three in ten depression cases and four in ten of any disorder, and higher cuts miss more—so at a single stage the SRQ-20 rules out better than it finds cases. Second, the criterion was a *reduced* CIDI covering a limited set of disorders (12-month prevalence of any assessed disorder only 3.8%, low by World Mental Health Survey standards); this narrow, episodic criterion both bounds the attainable AUC—the SRQ indexes current non-psychotic distress, a broader and partly different construct—and mechanically inflates the negative predictive value. Accordingly, negative predictive value was high (.98–.99) but reflects the low prevalence rather than screen quality, whereas positive predictive value was low (.09–.25) at these prevalences (Trevethan, 2017; Leeflang et al., 2013). The practical implication is a substantial false-positive load at low cut-points, best handled by a two-stage design in which SRQ-positive respondents receive a brief confirmatory interview, rather than by abandoning the low cut (Table 4; Figure 4).

**Figure 4.**
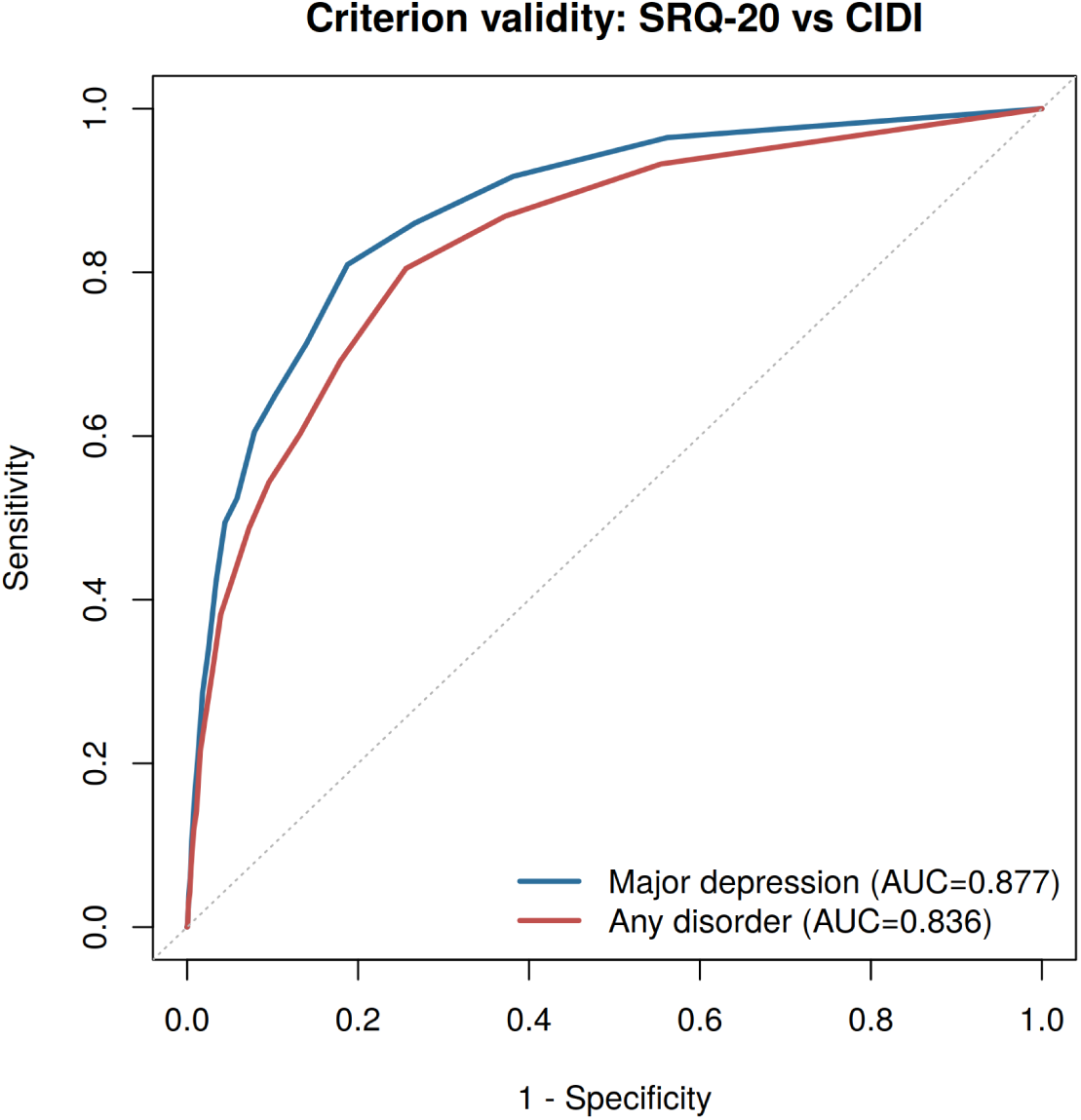
ROC curves of the SRQ-20 against 12-month CIDI major depression and any disorder.

**Table 4.**
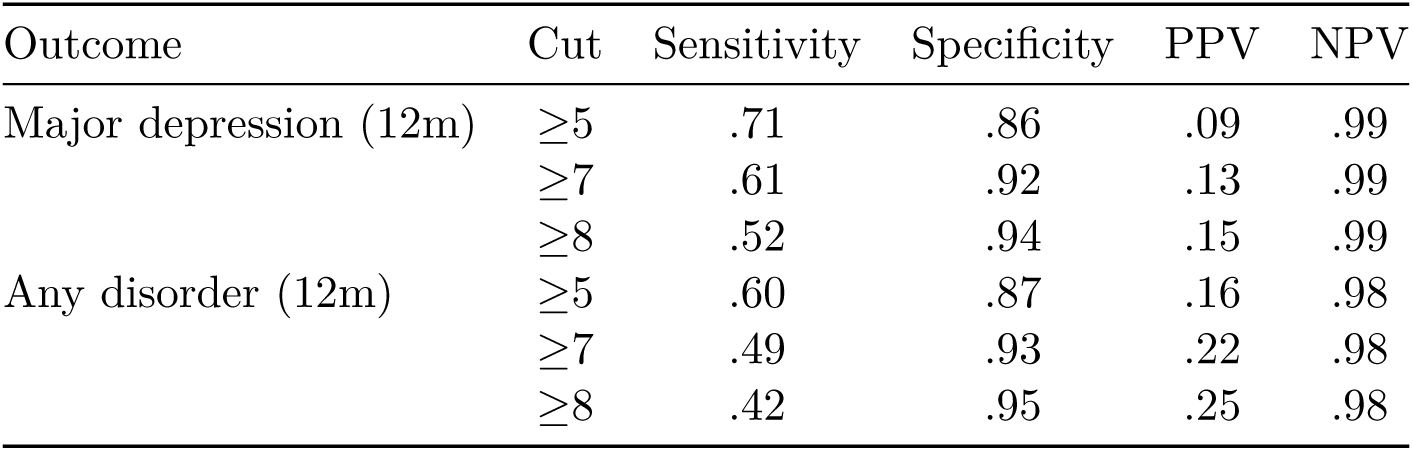
Design-weighted (FEX) criterion validity. Weighted AUC: major depression .877 (95% CI [.845, .900]); any disorder .836 ([.807, .859]). Youden-optimal cut (depression) *≥*4, stable under .632 correction (sensitivity .76*→*.75; specificity .79*→*.79). PPV = positive predictive value; NPV = negative predictive value.

| Outcome | Cut | Sensitivity | Specificity | PPV | NPV |
| --- | --- | --- | --- | --- | --- |
| Major depression (12m) | $\geq 5$ | .71 | .86 | .09 | .99 |
| | $\geq 7$ | .61 | .92 | .13 | .99 |
| | $\geq 8$ | .52 | .94 | .15 | .99 |
| Any disorder (12m) | $\geq 5$ | .60 | .87 | .16 | .98 |
| | $\geq 7$ | .49 | .93 | .22 | .98 |
| | $\geq 8$ | .42 | .95 | .25 | .98 |

### 3.8 The 25-item extension

Adding the four psychotic items did not yield a coherent subscale: their internal consistency was poor (*α* = .45), driven by the grandiosity item (“you are much more important than others think”), which was endorsed by 48.5% of respondents and discriminated poorly (a = .63); the remaining items discriminated acceptably (thought interference 1.98, hallucinations 1.69, persecutory ideation 1.38). The 24-item set remained essentially unidimensional (eigenvalue ratio 7.5), a distress+psychosis two-factor model improved fit only marginally (CFI .932 *→* .942), and the extended form remained equivalent by conflict exposure (ΔCFI = .0001). These items should therefore be interpreted cautiously and not as a validated psychosis subscale; the grandiosity item, in particular, performs poorly in this population. The 20-item form is recommended (Table 5).

**Table 5.** SRQ-25 psychotic-item IRT parameters and subscale reliability. Psychotic subscale (4 items): Cronbach *α* = .45; average inter-item r = .21.

| Item | Endorsement (%) | a | item-total r |
| --- | --- | --- | --- |
| Persecutory ideation | 21.5 | 1.38 | .37 |
| Grandiosity | 48.5 | 0.63 | .27 |
| Thought interference | 4.9 | 1.98 | .30 |
| Hearing voices | 3.3 | 1.69 | .22 |

## 4. Discussion

Across a nationally representative adult sample, the SRQ-20 measured common mental distress equivalently in adults exposed and not exposed to armed conflict, and across regions. Unlike a conventional “non-significant DIF” result, this conclusion rests on equivalence testing: once the groups’ true difference in distress was separated from item behaviour, the smallest DIF the design could resolve was *≈*.03 (SIDS), yet every item effect fell within a pre-specified small-effect band and the net test-level bias was only +0.12 points (Lakens, 2017; Lakens et al., 2018). The result is a clean dissociation between the *level* of distress and its *measurement*: exposed adults were more distressed (impact +0.29 SD, rising to +0.43 SD among the directly victimised), yet the instrument indexed that distress on the same metric (Bell et al., 2012; Cuartas Ricaurte et al., 2019). This supports comparing exposed and non-exposed adults *within a comparable population*—the ENSM’s Colombian adults, who share language, translation, and cultural frame. We stress the scope: the study did not compare a conflict setting with a non-conflict setting, or one country with another, and it does not license such cross-national comparisons; it tests, and supports, the within-population individual-exposure comparisons that this literature routinely makes but rarely validates for the instrument at national scale (Steel et al., 2009; Charlson et al., 2019).

The item-response analysis adds three points of substance. First, separating impact from bias was decisive for the headline result. The configural analysis produced a uniform, same-signed DIF pattern that would ordinarily be read as bias; freeing the group latent mean revealed it instead as impact—the exposed group’s higher distress—leaving negligible residual DIF. Reporting the raw multiple-group output would have misstated a true group difference as measurement bias; the uniform-sign pattern usefully diagnoses when this has happened. Second, allowing discriminations to vary is empirically justified (the 2PL clearly outperforms an equal-slope model), the 2PL fits well at both the global and item levels, and the only departures from local independence are a handful of content-redundant pairs (two digestion items, two tiredness items, work/enjoyment, worthlessness/uselessness) that mildly inflate apparent dimensionality without threatening the unidimensional treatment of the total score. Third, the well-known gap between the classical reliability of the SRQ-20 (*α* = .85) and its IRT marginal reliability (.65) is fully explained by the concentration of information above the population mean: at the screening thresholds, where decisions are actually made, conditional reliability is high (.93–.94). A single coefficient therefore understates the scale’s quality precisely where it matters and overstates it in the low range; the SRQ-20 is well suited to detecting elevated distress and deliberately uninformative below it.

Three nuances sharpen the applied message. First, sex is only partially invariant: a few somatic/affective symptoms function differentially, and the net 0.85-point differential test functioning means raw SRQ-20 sex comparisons are biased in women’s direction and should be adjusted (Paraventi et al., 2015; Burnette et al., 2024). The contrast with conflict is instructive. The two comparisons involve a *similar* true difference in distress (impact +0.29 SD for exposure, +0.25 SD for sex), yet the conflict comparison is essentially free of measurement bias (net DTF +0.12, no salient item) while the sex comparison carries real, systematic bias (+0.85)—a reminder that the magnitude of a true group difference is no guide to whether the instrument measures it fairly. Second, criterion validity against the CIDI is good but bounded by a narrow criterion and only marginally inflated by content overlap; the recommended cut-points are stable on internal validation, the low PPV is a prevalence phenomenon (Trevethan, 2017; Leeflang et al., 2013), and single-stage sensitivity is modest, so two-stage screening is the sensible deployment. Third, the four psychotic add-on items do not constitute a reliable subscale, and the grandiosity item performs especially poorly: endorsed by nearly half the sample, it behaves less like a psychotic-experience item than like an artefact of how it is phrased in the ENSM’s Spanish translation, where “more important than others think” can read as ordinary self-regard. Researchers using the SRQ in Colombia should rely on the 20-item form and treat the psychotic items, if used at all, as loose flags rather than a measured dimension.

Strengths include a large, nationally representative sample; a modern psychometric toolkit (tetrachoric dimensionality with local-dependence modelling, a fully diagnosed IRT model, effect-size DIF, and design-weighted ROC); and—central to the equivalence claim—the combination of equivalence testing with purified anchoring and a freely estimated group mean, which distinguishes true group differences from measurement bias rather than relying on a non-significant omnibus test.

### 4.1. Limitations

Several limitations qualify these conclusions. The public microdata provide expansion weights but not sampling clusters or strata; we therefore used weighted point estimates with region-stratified bootstrap CIs, which do not fully capture the multistage design effect. This caveat applies not only to the ROC but to the equivalence intervals: because narrower CIs bias toward *declaring* equivalence, we report intervals inflated for a plausible design effect (DEFF *≈* 2), and the item-level conclusions are unchanged under that inflation. Exposure was a single self-reported, retrospective binary; at 7.7% endorsement it likely captures a proximal subset and leaves some affected adults in the non-exposed group. Such non-differential misclassification attenuates DIF toward the band—working in favour of the null we report—so it is a genuine threat; the dose–response analysis mitigates it, because equivalence held (and impact grew) in the cleaner directly-victimised contrast, which misclassification cannot easily explain away. The criterion was a reduced CIDI covering a limited set of episodic disorders, bounding the attainable AUC and inflating NPV. The mild local dependence among content-redundant item pairs, while immaterial to the total-score model, would matter for scoring narrow somatic or affective subscales. The exposed subgroup, adequate for the equivalence test of item effect sizes, is small for the exploratory network moderation (hence its supplementary status). Finally, the data are cross-sectional, so longitudinal invariance was not assessed.

### 4.2. Conclusions

The SRQ-20 is an essentially unidimensional, well-fitting, reliable, criterion-valid screen that functions equivalently across armed-conflict exposure—including among directly victimised adults—and across regions within the Colombian adult population, with only partial invariance by sex (a systematic bias of about 0.85 points on the 0–20 scale). It supports comparisons of exposed and non-exposed individuals within a comparable population when the 20-item form is used and the sex differential is acknowledged; on this evidence alone it does not license cross-national comparison.

## Data Availability

All data produced are available online. This study is a secondary analysis of the openly available public-use microdata of the 2015 Colombian National Mental Health Survey (ENSM 2015), available from Datos Abiertos Colombia (https://www.datos.gov.co/Salud-y-Protecci-n-Social/Salud-Mental/d53j-hn85). The de-identified analytic dataset derived from these microdata and all reproducible analysis code, which reproduce every table and figure in the manuscript, are openly available at https://github.com/pedrovelezpardo/srq20-ensm2015-validation

https://github.com/pedrovelezpardo/srq20-ensm2015-validation

## Declarations

### Ethics

Secondary analysis of de-identified public microdata from the ENSM 2015; the original survey received ethics approval and informed consent (Gómez-Restrepo et al., 2016a).

### Data availability

The ENSM 2015 microdata are publicly available from the Colombian Ministry of Health (Datos Abiertos / MinSalud). All analysis code and the derived analytic dataset that reproduce every table and figure are openly available at https://github.com/pedrovelezpardo/srq20-ensm2015-validation.

### Author contributions (CRediT)

Pedro Vélez-Pardo: Conceptualization, Methodology, Software, Formal analysis, Data curation, Validation, Visualization, Writing – original draft, Writing – review & editing, Supervision, Project administration. Daniela Sánchez Acosta: Conceptualization, Investigation, Validation, Writing – review & editing. Nadia Semenova Moratto-Vásquez: Writing – review & editing. Johana Marcela Quintero-Hoyos: Investigation, Resources, Writing – review & editing.

### Declaration of generative AI and AI-assisted technologies in the manuscript preparation process

During the preparation of this work the authors used Claude (Anthropic) to assist with reviewing the analysis code and with drafting and editing the manuscript. After using this tool, the authors reviewed and edited the content as needed and take full responsibility for the content of the published article.

