## Supplementary material for "Measurement equivalence of the SRQ-20 across armed-conflict exposure, sex, and region in Colombia (ENSM 2015)"

---

### Supplementary Methods

**S1. Dimensionality (tetrachoric).** Because items are dichotomous, dimensionality was assessed on the matrix of tetrachoric correlations. In addition to Horn's parallel analysis (which over-extracts with dichotomous items in very large samples), we report the ratio of the first to second eigenvalue and three bifactor-based indices—explained common variance (ECV = .71), $\omega$-hierarchical (.79), and the proportion of uncontaminated correlations (PUC = .70)—computed from a Schmid–Leiman transformation of an exploratory bifactor solution (`psych::omega`), which is distinct from the confirmatory bifactor model (the latter did not converge and is treated as uninformative rather than as evidence about specific factors). At a moderate PUC, an ECV near .71 with $\omega$-hierarchical near .80 indicates that a general factor dominates. One-factor, correlated two-factor, and (attempted) bifactor confirmatory models were estimated with WLSMV. Because a few content-redundant pairs inflate apparent multidimensionality, we additionally fitted a one-factor model with correlated residuals for the four locally dependent pairs (a testlet parameterisation): this raised fit from CFI = .945/RMSEA = .043/SRMR = .066 to CFI = .971/RMSEA = .032/SRMR = .053, closing most of the gap to the two-factor model (CFI = .957) without invoking a second substantive dimension.

**S2. Item-response model and diagnostics.** A unidimensional two-parameter logistic (2PL) model was fitted by marginal maximum likelihood. To justify allowing discriminations to vary, the 2PL was compared with a one-parameter (equal-slope) model by likelihood-ratio test and by AIC/BIC. Global model–data fit was evaluated with the limited-information M2* statistic and its RMSEA and SRMSR; item-level fit with the S-$X^2$ statistic and its RMSEA (Supplementary Table S5); and the local-independence assumption with Yen's Q3 residual correlations, flagging item pairs whose Q3 exceeded the mean Q3 by more than 0.20 (Supplementary Table S6). We report the discrimination (a) and difficulty (b) parameters, the test-information and standard-error functions, IRT marginal and empirical reliability, and the conditional reliability at the screening cut-points.

**S3. Differential item and test functioning (DIF/DTF), purified estimation.** Given the very large sample, in which negligible differences reach statistical significance, DIF was quantified with effect sizes rather than significance tests, using the signed expected-score difference (SIDS) from `mirt::empirical_ES` (expressed in expected-endorsement units, 0–1). The critical step is to separate genuine item bias from a group difference in the latent trait itself (*impact*): if a more-distressed focal group has its latent mean fixed at zero, that true difference is absorbed into the item parameters and appears as spurious, uniformly-signed DIF. We therefore used a **purified-anchor procedure**: an initial configural model ranked items by |SIDS|; the four most invariant items were fixed as anchors; and the model was refitted with the focal group's latent **mean and variance freely estimated**. SIDS were recomputed from this purified model, and the estimated focal-group latent mean is reported as the impact. In the armed-conflict contrast, purification changed the picture qualitatively: the configural SIDS were uniformly signed (all 20 items, net -0.6), the signature of impact leakage, whereas the purified SIDS were small and mixed in sign (13 positive, 3 negative; maximum |SIDS| = .033), with an estimated impact of +0.29 SD. Item-level equivalence was declared when a SIDS fell within a pre-specified band of |SIDS| < 0.10 (less than half a point on the 0–20 scale, i.e., under a ten-percentage-point difference in endorsement probability at equal latent distress). Each SIDS was bootstrapped (stratified resampling) for a 95% CI, and—because the public microdata omit the sampling design—intervals inflated for a plausible design effect (DEFF $\approx$ 2) are also reported. As a detectability check (not a formal equivalence-power calculation), the smallest resolvable item DIF was approximated as 2.8 $\times$ bootstrap SE. The net scale-level consequence is summarised as differential test functioning (DTF): the signed test difference (STDS, the sum of the signed item expected-score differences) and the unsigned test difference (UTDS, the sum of the **absolute** item expected-score differences), following Meade (2010); the DTF was bootstrapped for a 95% CI. Omnibus invariance used the change in the C2-based CFI between configural, metric, and scalar multiple-group 2PL models ($\Delta$CFI $\leq$ .010 indicating invariance).

**S3b. Severity-graded (dose–response) sensitivity.** To bound the effect of non-differential exposure misclassification (which attenuates DIF toward the equivalence band), the conflict DIF was repeated contrasting adults reporting **direct** victimisation (n = 286) with the non-exposed (n = 10,027), a cleaner and more severe exposure gradient. Impact rose to +0.43 SD, yet DIF remained within the band (maximum |SIDS| = .071, no item $\geq$ .10; signed/unsigned DTF = 0.27/0.35): equivalence holds, and strengthens, as exposure severity increases.

**S3c. Design-weighted sensitivity.** To check that unweighted calibration does not distort the item-level results, all 2PL parameters were re-estimated with the expansion weight (FEX; `mirt survey.weights`). Discriminations and difficulties were essentially unchanged (a: r = .99, maximum $|\Delta a|$ = .15; b: r = .99, maximum $|\Delta b|$ = .11), so the DIF and dimensionality results are not artefacts of ignoring the sampling weights.

**S4. Psychometric-network analysis (exploratory).** The symptom network (Figure S1) was estimated as an Ising model via $\ell$1-regularised logistic neighbourhood selection (eLasso; `IsingFit`), with the EBIC hyperparameter $\gamma = 0.25$, and visualised with `qgraph`. Network stability was verified by case-dropping subset bootstrap (1,000 resamples; `bootnet`): the centrality-stability (CS) coefficient for strength and expected influence was 0.75 (> 0.50), indicating interpretable centrality. Structural invariance by conflict exposure was examined with a moderated network model (mixed graphical model; `mgm`) in which armed-conflict exposure moderated all symptom–symptom edges on the full sample. The model retained 115 pairwise edges and zero moderation effects, so the networks conditioned on exposure were indistinguishable (Figure S2). This analysis is presented as **exploratory and power-limited**: with roughly 838 exposed respondents, an $\ell$1-regularised estimator has limited power to recover moderation effects, so "zero moderated edges" is as much the expected output of an under-powered regularised model as evidence of true structural invariance. It corroborates but does not independently establish the equivalence result, which rests entirely on the purified equivalence-tested item-response analysis in the main text. A split-sample network-comparison test (`NetworkComparisonTest`) was consistent once the large group-size asymmetry ($\approx$838 exposed vs $\approx$10,000 non-exposed) was taken into account; the apparently sparser exposed network in a naïve split reflects sample-size–dependent regularisation rather than a true structural difference (at matched n the difference in global strength disappears).

### Supplementary Tables

**Table S1.** Exploratory factor analysis (tetrachoric correlations, two-factor oblimin): item loadings.

| Item | Factor 1 (somatic) | Factor 2 (psychological) |
|---|---|---|
| headaches | 0.764 | -0.239 |
| poor appetite | 0.652 | -0.028 |
| sleep badly | 0.683 | 0.022 |

| Item | Factor 1 (somatic) | Factor 2 (psychological) |
|---|---|---|
| easily frightened | 0.715 | -0.086 |
| hands shake | 0.42 | 0.215 |
| nervous/tense | 0.7 | 0.155 |
| poor digestion | 0.639 | -0.043 |
| trouble thinking | 0.419 | 0.385 |
| unhappy | 0.596 | 0.265 |
| cry more | 0.685 | 0.06 |
| little enjoyment | 0.243 | 0.669 |
| indecisive | 0.319 | 0.502 |
| work suffering | 0.014 | 0.836 |
| no useful role | -0.112 | 0.846 |
| lost interest | 0.38 | 0.481 |
| worthless | 0.062 | 0.666 |
| suicidal ideation | 0.436 | 0.224 |
| tired all the time | 0.459 | 0.373 |
| stomach discomfort | 0.605 | 0.125 |
| easily tired | 0.481 | 0.34 |

**Table S2.** One-factor confirmatory factor analysis (WLSMV): standardized loadings. All loadings p < .001.

| Item | Std. loading | SE | p |
|---|---|---|---|
| headaches | 0.521 | 0.013 | <0.001 |
| poor appetite | 0.596 | 0.015 | <0.001 |
| sleep badly | 0.667 | 0.012 | <0.001 |
| easily frightened | 0.613 | 0.011 | <0.001 |
| hands shake | 0.585 | 0.017 | <0.001 |
| nervous/tense | 0.815 | 0.008 | <0.001 |
| poor digestion | 0.591 | 0.015 | <0.001 |
| trouble thinking | 0.739 | 0.012 | <0.001 |
| unhappy | 0.827 | 0.008 | <0.001 |
| cry more | 0.735 | 0.011 | <0.001 |
| little enjoyment | 0.834 | 0.01 | <0.001 |
| indecisive | 0.747 | 0.011 | <0.001 |
| work suffering | 0.776 | 0.013 | <0.001 |
| no useful role | 0.639 | 0.022 | <0.001 |
| lost interest | 0.773 | 0.011 | <0.001 |
| worthless | 0.64 | 0.021 | <0.001 |
| suicidal ideation | 0.612 | 0.017 | <0.001 |
| tired all the time | 0.796 | 0.01 | <0.001 |
| stomach discomfort | 0.698 | 0.014 | <0.001 |
| easily tired | 0.779 | 0.009 | <0.001 |

**Table S3.** Purified vs configural item-level DIF by armed-conflict exposure, with anchor items flagged. Every purified |SIDS| < .10 (maximum .033).

| Item | SIDS (purified) | SIDS (configural) | Anchor |
|---|---|---|---|
| suicidal ideation | 0.033 | 0.047 | — |
| easily frightened | -0.023 | 0.026 | — |
| headaches | 0.018 | 0.058 | — |
| hands shake | 0.014 | 0.027 | — |
| easily tired | 0.014 | 0.057 | — |
| poor appetite | 0.012 | 0.033 | — |
| unhappy | 0.012 | 0.053 | — |
| work suffering | 0.011 | 0.023 | — |
| stomach discomfort | 0.01 | 0.029 | — |
| poor digestion | 0.009 | 0.03 | — |
| cry more | 0.008 | 0.034 | — |
| nervous/tense | -0.007 | 0.033 | — |
| little enjoyment | 0.005 | 0.021 | — |
| sleep badly | 0.003 | 0.039 | — |
| indecisive | -0.002 | 0.021 | — |
| trouble thinking | 0.001 | 0.023 | — |
| no useful role | 0 | 0.003 | Yes |
| lost interest | 0 | 0.021 | Yes |
| worthless | 0 | 0.02 | Yes |
| tired all the time | 0 | 0.018 | Yes |

**Table S4.** Purified vs configural item-level DIF by sex. Three items exceed the ±0.10 band (easily frightened .18, headaches .10, cry more .10).

| Item | SIDS (purified) | SIDS (configural) | Anchor |
|---|---|---|---|
| easily frightened | 0.181 | 0.2 | — |
| headaches | 0.104 | 0.126 | — |
| cry more | 0.1 | 0.107 | — |
| unhappy | 0.075 | 0.094 | — |
| poor digestion | 0.056 | 0.066 | — |
| easily tired | 0.055 | 0.078 | — |
| nervous/tense | 0.049 | 0.07 | — |
| poor appetite | 0.044 | 0.055 | — |
| sleep badly | 0.035 | 0.057 | — |
| tired all the time | 0.035 | 0.047 | — |
| stomach discomfort | 0.031 | 0.04 | — |
| indecisive | 0.029 | 0.041 | — |
| little enjoyment | 0.017 | 0.024 | — |
| trouble thinking | 0.015 | 0.028 | — |
| lost interest | 0.013 | 0.025 | — |
| suicidal ideation | 0.009 | 0.018 | — |
| hands shake | 0 | 0.009 | Yes |
| work suffering | 0 | 0.005 | Yes |
| no useful role | 0 | 0.002 | Yes |
| worthless | 0 | 0.009 | Yes |

**Table S5.** Item fit (S-$X^2$) for the 2PL model. All items adequate in magnitude (fit RMSEA $\leq .012$); five flagged at $p < .05$ given the large sample.

| Item | S-$X^2$ | df | RMSEA | p | Fit |
|---|---|---|---|---|---|
| headaches | 12.71 | 15 | 0.0 | 0.624 | adequate |
| poor appetite | 10.21 | 16 | 0.0 | 0.856 | adequate |
| sleep badly | 5.48 | 15 | 0.0 | 0.987 | adequate |
| easily frightened | 12.01 | 15 | 0.0 | 0.678 | adequate |
| hands shake | 14.92 | 16 | 0.0 | 0.531 | adequate |
| nervous/tense | 12.77 | 13 | 0.0 | 0.466 | adequate |
| poor digestion | 28.58 | 16 | 0.009 | 0.027 | trivial misfit |
| trouble thinking | 14.12 | 15 | 0.0 | 0.517 | adequate |
| unhappy | 15.82 | 13 | 0.004 | 0.259 | adequate |
| cry more | 28.83 | 15 | 0.009 | 0.017 | trivial misfit |
| little enjoyment | 18.03 | 15 | 0.004 | 0.261 | adequate |
| indecisive | 17.95 | 15 | 0.004 | 0.265 | adequate |
| work suffering | 8.73 | 15 | 0.0 | 0.891 | adequate |
| no useful role | 13.99 | 16 | 0.0 | 0.599 | adequate |
| lost interest | 19.96 | 15 | 0.006 | 0.174 | adequate |
| worthless | 40.76 | 16 | 0.012 | 0.001 | trivial misfit |
| suicidal ideation | 20.97 | 16 | 0.005 | 0.18 | adequate |
| tired all the time | 33.85 | 15 | 0.011 | 0.004 | trivial misfit |
| stomach discomfort | 21.0 | 15 | 0.006 | 0.137 | adequate |
| easily tired | 26.16 | 14 | 0.009 | 0.025 | trivial misfit |

**Table S6.** Local dependence (Yen's Q3), top item pairs. Four content-redundant pairs exceed the criterion ($|Q3 - mean| > .20$).

| Item i | Item j | Q3 | LD |
|---|---|---|---|
| work suffering | little enjoyment | 0.278 | LD |
| stomach discomfort | poor digestion | 0.265 | LD |
| easily tired | tired all the time | 0.22 | LD |
| easily tired | unhappy | -0.202 | — |
| easily tired | nervous/tense | -0.181 | — |
| worthless | no useful role | 0.168 | LD |
| little enjoyment | nervous/tense | -0.161 | — |
| easily tired | cry more | -0.154 | — |
| cry more | unhappy | 0.152 | — |
| work suffering | nervous/tense | -0.151 | — |
| stomach discomfort | unhappy | -0.145 | — |
| indecisive | trouble thinking | 0.142 | — |
| unhappy | poor digestion | -0.142 | — |
| work suffering | cry more | -0.14 | — |
| tired all the time | trouble thinking | -0.135 | — |

**Table S7.** Purified DIF for region (each macro-region vs the pooled remainder); region maximum $|SIDS| = .046$.

| Region | n | max | SIDS | |
|---|---|---|---|---|
| 1 | 2170 | 0.038 | 0.072 | 0 |
| 2 | 2360 | 0.025 | -0.248 | 0 |
| 3 | 1726 | 0.034 | 0.122 | 0 |
| 4 | 2577 | 0.019 | -0.007 | 0 |
| 5 | 2032 | 0.046 | -0.081 | 0 |

Test-level differential test functioning (DTF) summary across the purified contrasts (STDS = signed, UTDS = unsigned test difference):

| Contrast | STDS | UTDS | max | SIDS | |
|---|---|---|---|---|---|
| Armed-conflict exposure | 0.118 | 0.182 | 0.033 | 0 | 0.285 |
| Direct victimisation | 0.274 | 0.346 | 0.071 | 0 | 0.434 |
| Sex, women vs men | 0.849 | 0.849 | 0.181 | 3 | 0.245 |

### Supplementary Figures

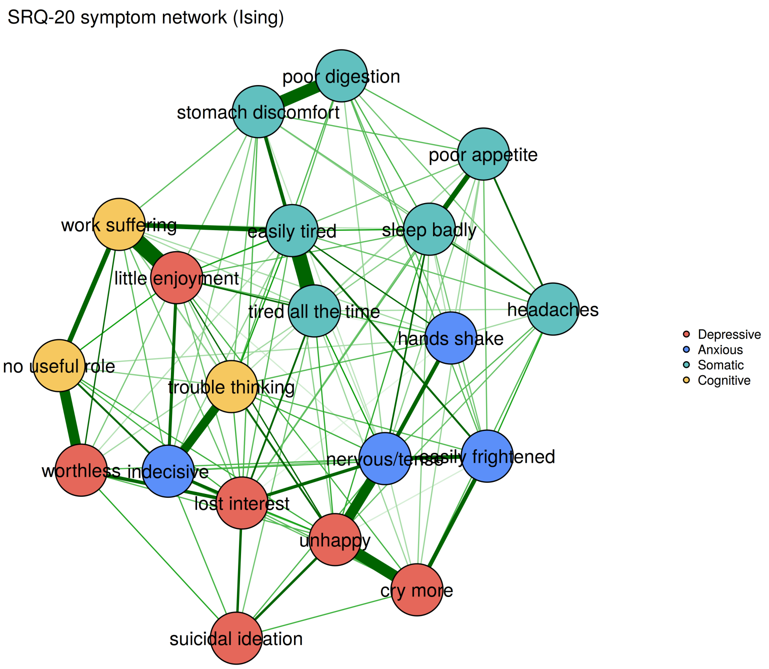


**Figure S1.** SRQ-20 symptom network (Ising model), full adult sample; nodes coloured by content domain.

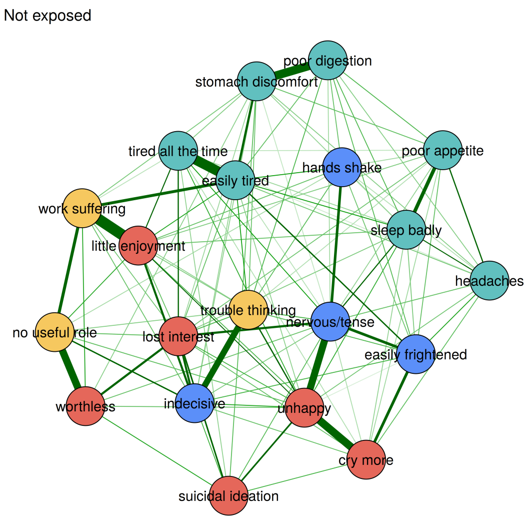


**Figure S2.** Symptom network conditioned on armed-conflict exposure (moderated network model); the two panels are indistinguishable (zero moderated edges), providing exploratory structural evidence.

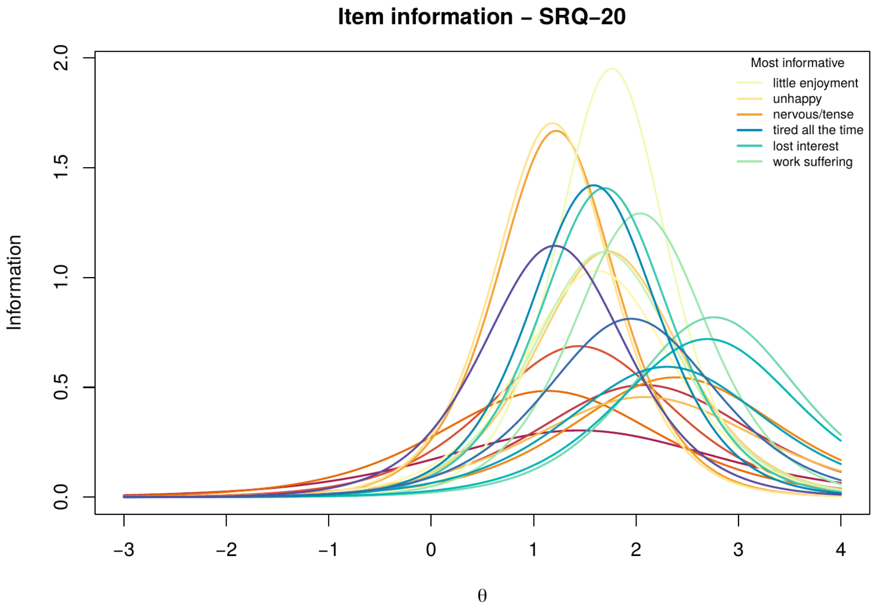


**Figure S3.** Item information curves, SRQ-20.

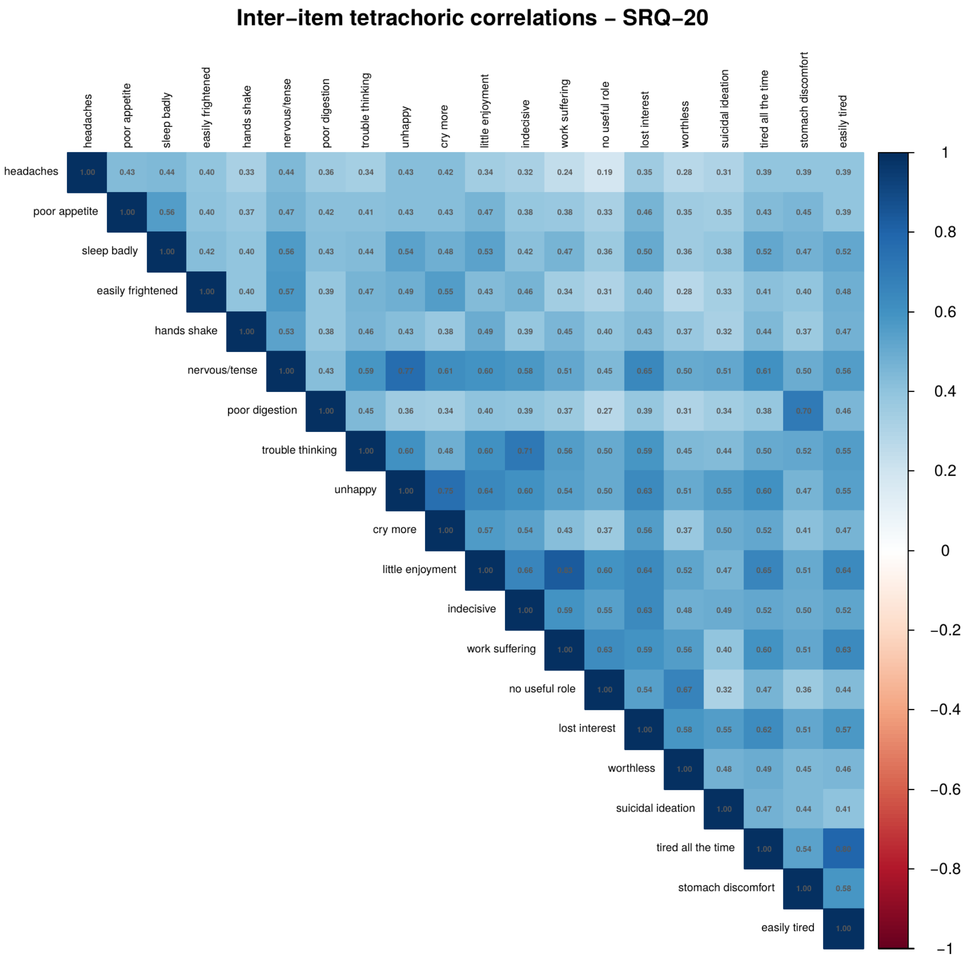


**Figure S4.** Inter-item tetrachoric correlation matrix, SRQ-20.

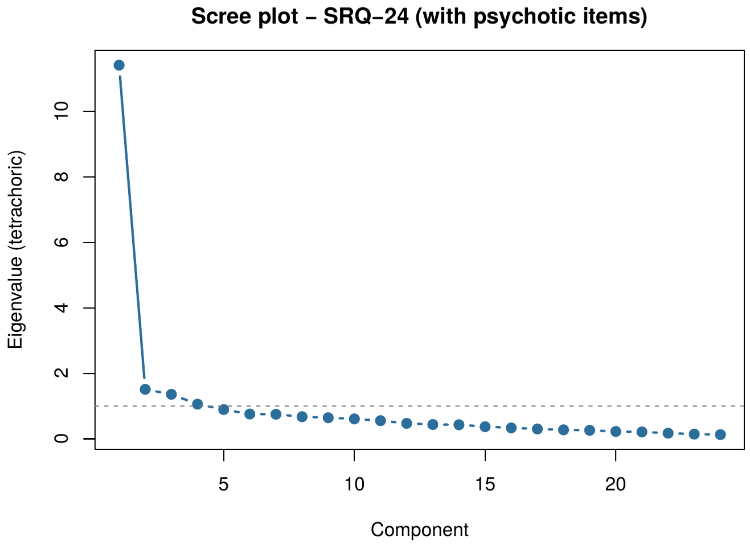


**Figure S5.** Scree plot of the 24-item extension including the four psychotic-experience items.

### Supplementary Software References

Epskamp, S., Borsboom, D., & Fried, E. I. (2018). Estimating psychological networks and their accuracy: A tutorial paper. *Behavior Research Methods, 50*(1), 195–212. https://doi.org/10.3758/s13428-017-0862-1

Epskamp, S., Cramer, A. O. J., Waldorp, L. J., Schmittmann, V. D., & Borsboom, D. (2012). qgraph: Network visualizations of relationships in psychometric data. *Journal of Statistical Software, 48*(4), 1–18. https://doi.org/10.18637/jss.v048.i04

Haslbeck, J. M. B., & Waldorp, L. J. (2020). mgm: Estimating time-varying mixed graphical models in high-dimensional data. *Journal of Statistical Software, 93*(8), 1–46. https://doi.org/10.18637/jss.v093.i08

van Borkulo, C. D., & Epskamp, S. (2023). *IsingFit: Fitting Ising models using the eLasso method* (R package version 0.4) [Computer software]. https://CRAN.R-project.org/package=IsingFit

van Borkulo, C. D., van Bork, R., Boschloo, L., Kossakowski, J. J., Tio, P., Schoevers, R. A., Borsboom, D., & Waldorp, L. J. (2023). Comparing network structures on three aspects: A permutation test. *Psychological Methods, 28*(6), 1273–1285. https://doi.org/10.1037/met0000476
